# Prolonged viral shedding from noninfectious individuals confounds wastewater-based epidemiology

**DOI:** 10.1101/2023.06.08.23291144

**Authors:** Tin Phan, Samantha Brozak, Bruce Pell, Stanca M. Ciupe, Ruian Ke, Ruy M. Ribeiro, Anna Gitter, Kristina D. Mena, Alan S. Perelson, Yang Kuang, Fuqing Wu

## Abstract

Wastewater surveillance has been widely used to track and estimate SARS-CoV-2 incidence. While both infectious and recovered individuals shed virus into wastewater, epidemiological inferences using wastewater often only consider the viral contribution from the former group. Yet, the persistent shedding in the latter group could confound wastewater-based epidemiological inference, especially during the late stage of an outbreak when the recovered population outnumbers the infectious population. To determine the impact of recovered individuals’ viral shedding on the utility of wastewater surveillance, we develop a quantitative framework that incorporates population-level viral shedding dynamics, measured viral RNA in wastewater, and an epidemic dynamic model. We find that the viral shedding from the recovered population can become higher than the infectious population after the transmission peak, which leads to a decrease in the correlation between wastewater viral RNA and case report data. Furthermore, the inclusion of recovered individuals’ viral shedding into the model predicts earlier transmission dynamics and slower decreasing trends in wastewater viral RNA. The prolonged viral shedding also induces a potential delay in the detection of new variants due to the time needed to generate enough new cases for a significant viral signal in an environment dominated by virus shed by the recovered population. This effect is most prominent toward the end of an outbreak and is greatly affected by both the recovered individuals’ shedding rate and shedding duration. Our results suggest that the inclusion of viral shedding from non-infectious recovered individuals into wastewater surveillance research is important for precision epidemiology.

## Introduction

SARS-CoV-2 concentration in wastewater is strongly correlated to case report data (*R* > 0.7) (1), supporting the effort of using wastewater-based epidemiology (WBE) to estimate infection incidence (2, 3). However, gastrointestinal tract viral shedding exhibits great heterogeneity among individuals, and a significant portion of clinically recovered individuals (i.e., individuals who are no longer infectious) shed virus in fecal specimens for weeks to months after the infectious phase (4–7). As an outbreak progresses, the number of recovered individuals accumulates, which results in an increase in their contribution to the overall viral RNA load in wastewater. While the prolonged shedding of SARS-CoV-2 in feces is well documented (4–6), to what extent the persistent shedding impacts the epidemiological inference of wastewater data remains unexplored. Based on recent studies, we propose the integration of wastewater data with within-host (viral shedding dynamics) and between-host (transmission dynamics) models to demultiplex the viral shedding from infectious and recovered phases (8, 9). The central idea is to use a population-level viral shedding profile, segmented according to the infection state, to connect viral RNA in wastewater with standard transmission dynamics models (SI Appendix). Our goal is to examine the potential impact of the virus shed by the recovered population on the capability of WBE to capture the epidemic landscape.

## Results and Discussion

### Viral shedding by the recovered population alters the correlation of virus measured in wastewater and case report data

We fit a Susceptible-Exposed-Infectious-Recovered-Virus (SEIR-V) model (Eq. 1, SI Appendix) including a population viral shedding function (Eq. 5, SI Appendix) to viral RNA measurements in wastewater collected from the Greater Boston area from 10/01/2020 to 02/27/2021 (1, 3), under the assumption that individuals who stop shedding virus are removed from the recovered class. The time frame was chosen to limit the data to a single wave of the COVID-19 pandemic (Alpha variant B.1.1.7) when the vaccination rate was low in the population. For a proof-of principle demonstration, we assume that recovered individuals on average shed virus at 50% of the mean viral shedding rate of infectious individuals for 14 days. The model captures the viral dynamics in wastewater well and the inferred transmission dynamics reflect the trend of the reported incidence (Fig. 1A-B). The model-predicted transmission dynamics, however, precede the reported incidence by about 3 weeks (as compared by peak timing – vertical dashed lines in Fig. 1B). The model-inferred incidence exceeds the reported daily incidence by 3-5 times near the beginning of Oct. and at the end of Feb. and over 10 times around Dec. Both observations are consistent with the expected lead time of wastewater data relative to case report data and the underreported incidence rates (10, 11).

**Figure 1.**
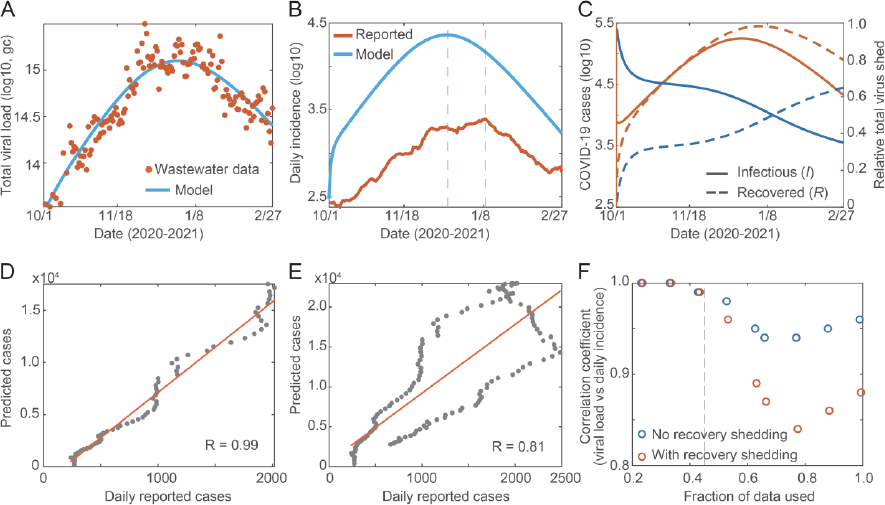
Viral shedding from the recovered population alters the correlation of virus measured in wastewater and case report data. Predicted viral dynamics obtained from fitting model Eq. 1 to viral shedding data of both infectious and recovered individuals. (**A**) Theoretical viral dynamics (blue curve) obtained by fitting the daily production of *V* in Eq. 1 to daily measurements of viral RNA (brown dots) in wastewater for the Greater Boston area. (**B**) Model-predicted daily incidence (blue line) vs. reported incidence (brown curve). The vertical dashed lines are the incidence peak time. (**C**) Left y-axis: model predicted number of infectious (solid brown curve) and recovered individuals (dotted brown curve). Right y-axis: relative contribution to the viral RNA measurements from the infectious (solid blue curve) and recovered (dotted blue curve) populations. (**D-E**) The correlation coefficient between predicted cases and reported cases decreases as more data are used for the fitting (within one wave). D only uses pre-peak data. E contains increasingly more data after the peak. (**F**) The model-predicted correlation between daily viral measurements in wastewater and daily incidence with and without the recovered shedding in the model. If the fraction of data used is 1, the data for one entire outbreak is used for the correlation. The vertical dashed line indicates the transmission peak.

Next, we compared the dynamics of the infectious and the recovered populations. As shown in Fig. 1C, the number of recovered individuals increases quickly and exceeds the infectious population. While the infectious population contributes most of the viral RNA in wastewater near the beginning of this epidemic wave, the recovered population eventually surpassed their viral contribution. Although these quantitative results may vary with different assumed viral shedding rates and shedding durations for the recovered population, they show how the viral shedding from the recovered population becomes more significant as an outbreak progresses.

The increase of virus shed by the recovered population creates a transient period where wastewater data may not directly reflect the incidence and dynamics of active infection. An intuitive approach to validate this inference is by comparing the correlation between wastewater viral measurements and reported incidence using data spanning different phases of an outbreak. During the early phase of an outbreak, wastewater data should be highly correlated with the reported daily new cases since the infectious population contributes substantially more virus to wastewater. However, the correlation should decrease as the outbreak progresses with the increase of the virus shed by the recovered population. We explicitly demonstrated this point by fitting the same model to increasingly larger data sets (Fig. 1D and 1E). We observe a clear decreasing trend in the correlation coefficient between the model-inferred daily incidence and the reported daily incidence with the inclusion of data post-transmission peak. Additionally, the model-simulated correlations (SI Appendix) between daily viral measurement and incidence show that the inclusion of viral shedding by the recovered population predicts a faster decline of the correlation coefficient as the outbreak progresses (Fig. 1F). Note that the increasing trend near the end is a result of the convergence of daily viral measurement in wastewater and incidence at the end of the outbreak. Thus, the exclusion of the virus shed by the recovered population is likely to have a small effect on the model-inferred transmission dynamics in the early phase of an outbreak (Fig. 1D and 1F). However, when using wastewater surveillance to track emerging cases toward the end of an outbreak or to connect multiple waves, it is necessary to include the virus shed by the recovered population to accurately capture transmission.

### Inclusion of recovered shedding predicts an earlier peak time of the outbreak

To better explain the mechanisms behind the changes in the correlation between wastewater data and case report data, we tested the impact of varied shedding rates and duration from the recovered individuals on model-inferred transmission dynamics. Fig. 2A shows the model fitting to viral load in wastewater under three scenarios: 1) no shedding from recovered individuals, 2) HRMD: high recovered shedding rate (75% of the infectious shedding rate) for a moderate duration of 14 days, 3) MRLD: moderate recovered shedding rate (50% of the infectious shedding rate) with a long duration of 21 days. Model fits under the three scenarios are similar, but the higher shedding rates and longer shedding durations result in slightly slower decreasing trends of the total viral load (Fig. 2A). A remarkable difference is observed in the daily incidence. The inclusion of viral shedding by the recovered population shifts the transmission peak by 6- and 10-day earlier under MRLD and HRMD scenarios, respectively (vertical dashed lines, Fig. 2B). These results suggest that the actual incidence peak time of the Alpha variant outbreak may be much earlier than the clinically reported cases and the change in the correlation is due in part to the differences in the viral shedding dynamics between the infectious and the recovered phases.

**Figure 2.**
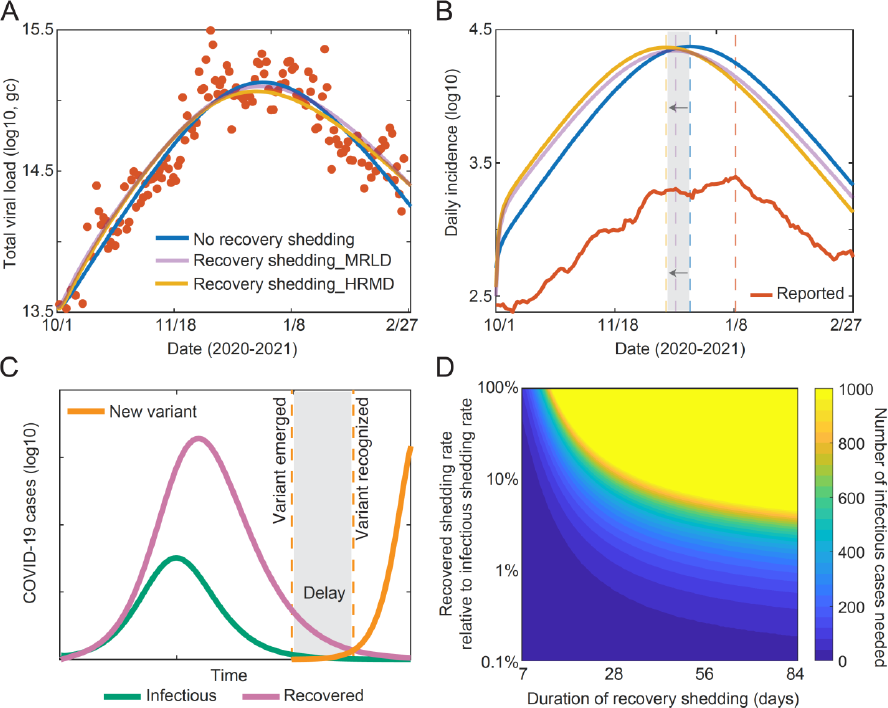
The inclusion of recovered shedding predicts an earlier peak time of the outbreak and a delay in the detection of an emerging variant by WBE. (**A**) Viral RNA in wastewater under different recovered shedding scenarios. Each model was fit to data from 10-01-2020 to 02-27-2021, which was chosen to restrict the outbreak to a single variant (brown data points). (**B**) Model inferred daily incidence under three recovered shedding scenarios. The vertical dashed curves are the corresponding peak times. (**C**) A schematic of tracking the emergence of a new variant near the end of an outbreak when most of the virus measured comes from the recovered population. (**D**) The number of new infectious cases needed to produce a sufficiently large viral signal, which depends on the recovered shedding rate and duration (Eqs. 9-11, SI Appendix). Warmer colors refer to a higher number of cases. This snapshot was taken when the outbreak reached 99.9% of its final size (as depicted by **C**) to show the potential of WBE to capture an emerging outbreak.

### The virus shed by the recovered population leads to a delay in the detection of an emerging variant by WBE

Several studies have looked at the minimum number of new cases required for the detection of virus in wastewater (12–14); however, none has looked into this issue in the presence of uncertainty due to the viral shedding from the recovered population. This is especially important during a transition period when a new variant is emerging near the end of an ongoing outbreak (Fig. 2C). The viral shedding from a large number of recovered individuals in wastewater may overshadow the viral shedding from new cases infected with the new variant, therefore undermining a rapid recognition of the emergence of the new variant based on the viral load in wastewater. We used the SEIR-V model to further quantify the relationship between shedding from recovered population and shedding from individuals with the new variant. Assuming the emerging variant has a similar viral shedding rate to its predecessor, we found that the number of required infectious cases with the new variant to signal an emerging outbreak is an increasing function of both the viral shedding rate and shedding duration of the recovered population (Fig. 2D, warmer colors). Since it takes time to generate the required number of infections for detection, this implies an increasing delay in the detection of an emerging outbreak as more new infections are needed. This delay is minimized if the viral shedding rate of the new variant is significantly higher than its predecessor (data not shown). Thus, tracking the viral shedding from the recovered population is important to mitigate the delay in the detection of new variants based on wastewater data.

The identification and mitigation of this time delay requires an accurate description of the population viral shedding profile, which may be variant-specific. Thus, integrating modeling analysis of routine wastewater surveillance data and genomic sequencing of wastewater samples (15) may better facilitate the identification of emerging variants circulating in the community. While our analysis focuses on the end of an outbreak to highlight the impact of prolonged shedding, similar analyses can be carried out to study the detection delay at different times during the outbreak and may need to account for the viral shedding from both the infectious and recovered population if they are not negligible.

In summary, we show that an understanding of the relative viral shedding during each phase of an infection is crucial to the implementation of precision WBE, which transpires beyond the study of the correlation between viral measurement in wastewater and case report data. Prolonged shedding can alter model-inferred transmission dynamics and lead to a delay in detection if not accounted for. Our analyses provide a proof-of-principle framework to quantify the impact of prolonged fecal shedding on wastewater-based estimation of new infections and the outbreak progression.

## Data Availability

All data produced in the present study are available upon reasonable request to the authors

## Materials and Methods

See SI Appendix.

## Data and Code Availability

The authors declare that the data and code will be provided with the publication of this study.

## Author Contributions

T.P., R.K., R.M.R., Y.K., and F.W. designed research; T.P., S.M., B.P., and F.W. performed research; T.P., S.M., R.K., R.M.R., K.D.M., A.S.P., Y.K., F.W. contributed new reagents/analytic tools; T.P., S.M., B.P., S.M.C., R.K., R.M.R., A.G., and F.W. analyzed data; and T.P., S.M., B.P., S.M.C., R.K., R.M.R., A.G., K.D.M., A.S.P., Y.K., and F.W. wrote the paper.

## Acknowledgments

This work is supported by Faculty Startup funding from the Center of Infectious Diseases at UTHealth, the UT system Rising STARs award, and the Texas Epidemic Public Health Institute (TEPHI) to F.W. This work was also supported by the Director’s postdoctoral fellowship at Los Alamos National Laboratory to T.P.; Y.K. and S.B. are partially supported by the US National Science Foundation Rules of Life program DEB -1930728 and the NIH grant 5R01GM131405-02.

## Supplementary Materials

### Methods

#### S1. SEIR-V model with temperature variation

We modified the SEIR-V model in Phan et al. (1) to include viral shedding from both the infectious and recovered populations. We also initially examined the viral shedding from the exposed class, which turned out to be negligible, due to the low shedding rate and short duration of the exposed class (not shown). Its implications are discussed in S6.

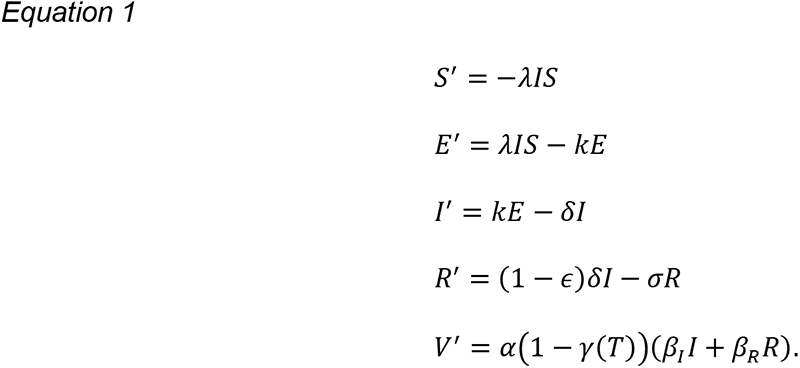

*S* is the susceptible population, *E* is the exposed population, *I* is the infectious population, and *R* is the recovered population that still sheds virus. Susceptible individuals are infected at a rate *λI*. Exposed individuals take on average 1/*k* days before becoming infectious. Infectious individuals recover or die after an average duration of 1/δ days. The disease-induced mortality rate is *ϵ*. However, since SARS-CoV-2-induced mortality is small, we set *ϵ* = 0 for simplicity. Recovered individuals shed virus for an average duration of 1/σ days. The cumulative viral RNA in wastewater (*V*) is contributed by both infectious and recovered individuals in this model at rates *β*_*I*_ and *β*_*R*_, respectively, as opposed to the model proposed by Phan et al. (1). α is the average fecal load per day. γ(*T*) is the fraction of virus lost in wastewater between shedding and collection (for measurement), which is assumed to be temperature-dependent (*T*). The temperature variation over the duration of the study is fitted to average daily temperature data (1) in Celsius.

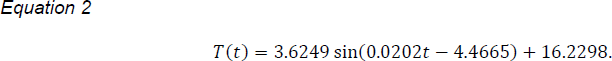

Where *t* is measured in days and *t* = 0 is 10/01/2020, see Fig. S1 in (1). The temperature-adjusted half-life is assumed to be an exponential decay of the form:

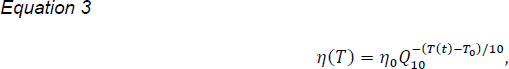

with η_0_ being the half-life (hours) at ambient temperature *T*_0_ and *Q*_10_ (= 2.5) being the temperature-dependent rate of change (1, 2). The temperature-dependent decay rate *ξ*(*T*) is approximated by a first-order decay function, which gives: 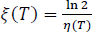. Assume viral RNA decays exponentially 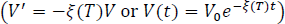, then the fraction of virus decay between shedding and collection is given by:

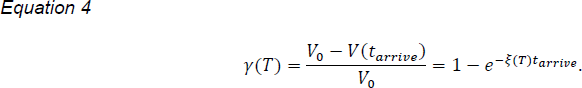

Here, *t_arrive_* is the time between shedding and collection, which we take to be 24 hours. The loss fraction γ is approximated between 0.03-0.28 for varying travel times and temperature, see SI in (1). However, this loss fraction could also be a function of the biochemical composition of the wastewater and microorganism community in the sewer. For example, an extremely harsh environment may result in a shorter viral half-life, which implies that the sample may be mostly influenced by viruses shed closer to the time of sampling.

Finally, we remark that *V*(*t*) is a proxy variable for the cumulative virus shed in wastewater. Its daily difference Δ*V*_*t*_, which represents the virus measured on day *t* is the quantity of interest, e.g., Δ*V*_*t*_ = *V*(*t*) − *V*(*t* − 1). Similarly, we estimate the daily incidence by looking at the cumulative number of infectious individuals. This is done by creating another proxy variable *C*(*t*) with *C*^′^(*t*) = *kE* and taking the daily difference Δ*C*_*t*_ to be the daily incidence on day *t*, e.g., Δ*C*_*t*_ = *C*(*t*) − *C*(*t* − 1). Note that while Δ*C*_*t*_ is a good approximation, it does not perfectly correspond to the daily report data, which varies based on many factors such as report time and symptom onset.

#### S2. The population level viral shedding function

The viral dynamics within an infected individual is relatively well defined (3, 4) and can be described by standard models of viral dynamics (5). Here, to simplify, we describe the viral levels in the gastrointestinal tract, which is proportional to the viral shedding rate, using a phenomenological function introduced by Phan et al. (1):

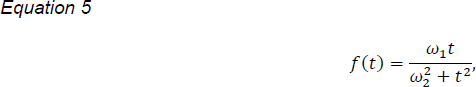

which peaks at 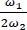 when *t* = *ω*_2_ Here. *ω*_1_ (log10 viral RNA copy per g) is a magnitude modifier and *ω*_2_ (day) influences the timing and magnitude of the viral shedding peak. Note that the viral shedding rate represents the viral concentration (per g of fecal matter) per day. This function has been shown to be a good approximation for the viral shedding dynamics (1). With this function, the total shedding between times *t*_1_ and *t*_2_ is given by

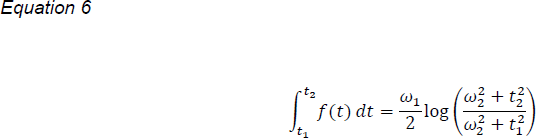

Where 0 ≤ *t*_1_ < *t*_2_ and log denotes the natural logarithm. We remark that the unit of *f*(*t*) is log10 viral RNA copy per g per day, meaning the integral ∫ *f*(*t*)*dt* has unit of log10 viral RNA copy per g.

The average exposed *E*, infectious *I*, and recovered with shedding *R* phases of an infection are classified based on the time since infection (S1). Thus, we can define *t*_*E*_, *t*_*I*_, and *t*_*R*_ (or 1/*k*, 1/*δ*, and 1/*σ*) to be the average time an infected person spent in these infection phases, respectively. The average viral shedding rate during the infectious phase *β*_*I*_ can be determined by calculating the average viral shedding over the infectious duration:

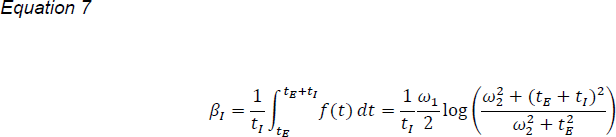

Note that the unit of *β*_*I*_ is log10 viral RNA copy per g per day. Based on the best-fit to the viral shedding data (1), we take *ω*_1_ = 71.97 log10 viral RNA copy per g and *ω*_2_ = 4 days. The estimated value of *β*_*I*_ with *t*_*E*_ = 3 days and *t*_*I*_ = 8 days is 7.65 log10 viral RNA copy per g per day. The viral shedding rate for the recovered population *β*_*R*_ is assumed to be a fraction of *β*_*I*_. For instance, the example in Fig. 1 assumes *β*_*R*_ = 0.5 × *β*_*I*_ and *t*_*R*_ = 14 days.

#### S3. Data fitting

When fitting the SEIR-V model to the viral RNA data in wastewater, we minimize the difference (*S_SE_*) between the measurement collected every 24-hour and the total virus produced at each time data point. Here, ^*V̂*^^(*t*_*d*_) is the total virus (e.g., viral RNA concentration × total flow) measured on day *t*_*d*_, where *t*_*d*_ is the time of the measurement. *V*(*t*), as given by Eq. 1, is a proxy of the cumulative viral RNA in wastewater. Thus, the viral RNA produced daily, given by 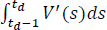, or Δ*V*_*t*_, is the comparable quantity to the viral RNA measured every 24-hour.

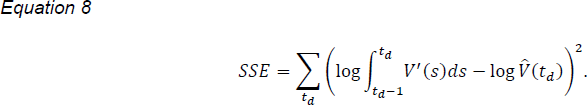

The computational minimization procedure is done using the function *fmincon* and *multistart* (500 random initial guesses) in MATLAB.

The best-fit in Fig. 1A gives *λ* = 9.36 × 10^−8^ per day per person, *α*, = 126 g, and *E*(0) = 145 people. Note that the initial for the infected population is approximated by 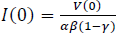, where *V*(0) is taken as the first viral RNA measurement and *E*(0) and *R*(0) are assumed to be 0. The parameters *k* and *δ* are fixed to 1/3 and 1/8 per day, respectively (3), and *σ* is fixed to 1/14 per day for the scenario in Fig. 1.

#### S4. Theoretical correlation between daily viral measurement and incidence with and without recovered shedding

We use the model to demonstrate how recovered shedding may affect the correlation between viral measurements and incidence. The methods that are often used to relate daily viral measurements to incidence data vary in literature (6, 7). However, here, we will simply look at the correlation of daily viral measurement (Δ*V*_*t*_) and incidence (Δ*C*_*t*_). To produce Fig. 1I, we use the best fit parameters for Fig. 1A (given in S3). The model without recovered shedding has *β*_*R*_ = 0.

#### S5. Simulation study of the impact of recovered shedding on the capability of WBE

One aspect of our study is to understand how the population average viral shedding profile affects the sensitivity of wastewater surveillance data to track new cases over the course of an outbreak. Since the viral shedding function *f*(*t*) is fixed in structure and was fitted to only one data set, it may not be able to fully capture the true dynamics of viral shedding. Thus, we want to examine how variations in *β*_*R*_ and *σ* (or 1/*t*_*R*_) affect the model inferred transmission dynamics. To focus our analysis on the impact of viral shedding from the recovered individuals, we vary only the recovered shedding rate *β*_*R*_ and the duration *σ* while keeping all other parameters in the model (Eq. 1) fixed as follow: *k* = 1/3 per day, *δ* =1/8 per day, *λ* = 4 × 10^−7^per person per day, *α* = 125 g, *γ* = 0.1, *β*_*I*_ = 7.65 log10 viral RNA copy per g per day, *S*(0) = 1 × 10^6^ people, *E*(0) = 1 person, *I*(0) = 1 person, and *R*(0) = 1 person.

The net virus shed by the *R* class varies over the course of an outbreak. Here, we limit the analysis to the end of an outbreak, where virus shed by the recovered population contributes close to 100% of viral measurements. In this scenario, to detect a significant change in the virus in wastewater, we need enough new infections (or infectious cases) that can generate at least 5% (assumed) of the viral shed by the recovered population on day *t*_*d*_ denoted by

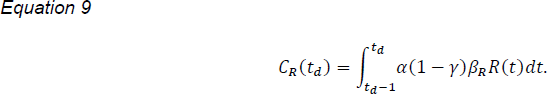

Let *N*(*t*_*d*_) be the minimum number of new infections needed to match the viral signal from the recovered population, then *N*(*t*_*d*_) must satisfy:

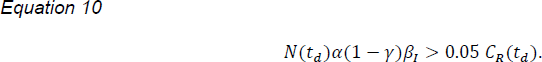

Solving for N, we have

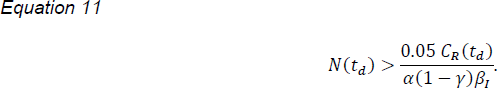

For reporting purposes (Fig. 2C-D), we round up *N* to the nearest integer. The heat map (Fig. 2D) is generated by calculating *N* for the average recovered shedding rate *β*_*R*_ varying between 0.001 to 1 of *β*_*I*_ and the average recovered shedding duration *σ* varying between 1/7 to 1/84 per day (or an average recovered shedding duration from 1 week to 12 weeks). The relative time (99.9%) of the outbreak is defined as the time when the cumulative recovered population given by 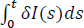 reaches 99.9% of its final size 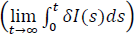.

#### S6. Further considerations

Our analyses ignore an intrinsic detection delay with WBE due to the low viral shedding of the exposed phase. Exposed individuals likely shed virus at a very low rate compared to infectious and recovered individuals. Thus, it is logical to assume that WBE is unlikely to detect the virus shed by an exposed individual during an ongoing outbreak, leading to an intrinsic detection delay of new cases using WBE. This detection delay may not be longer than 5 days, which is the average time from infection to symptom onset, signaling high respiratory tract viral load (3). This issue could be studied in detail if an accurate population level viral shedding dynamics is available. However, to our knowledge, there has not been reported data on virus shed from the gastrointestinal tract during the exposed phase of infection. Thus, even the population level viral shedding function *f*(*t*) used here may not accurately reflect the early viral shedding dynamics.

Furthermore, positive detection of SARS-CoV-2 virus was possible in only about 50% of samples from infected individuals (8, 9), which suggests the possibility of gastrointestinal tract infection being an inherent stochastic event. These important factors, among others, were not considered in our study due to limited data but should be analyzed further to better understand and improve the utility of WBE.

